# Neurophysiological signatures of Stanford Neuromodulation Therapy in treatment resistant depression

**DOI:** 10.1101/2025.08.13.25333640

**Authors:** Davide Momi, Derrick Buchanan, Masataka Wada, Andrew D. Geoly, Noriah J. Johnson, Eleanor J Cole, Adi Maron-Katz, Claudia Tischler, Christopher C. Cline, Manish Saggar, Daphne Voineskos, John D. Griffiths, Camarin E. Rolle, Corey J. Keller, Nolan Williams

## Abstract

**Objective:** Stanford Neuromodulation Therapy (SNT) has demonstrated impressive efficacy for treatment-resistant depression (TRD), but its neurophysiological mechanisms remain poorly understood. This study investigated the neurophysiological correlates of SNT using transcranial magnetic stimulation with electroencephalography (TMS-EEG).

**Method:** A double-blind, randomized, sham-controlled clinical trial was conducted at Stanford University. Twenty-four participants with TRD (12 active, 12 sham) received TMS-EEG measurements at baseline, before/after each treatment day across the 5-day protocol, and at 1-month follow-up. The intervention consisted of active or sham SNT protocol (10 daily sessions of intermittent theta-burst stimulation to the left dorsolateral prefrontal cortex over 5 consecutive days). Main outcome measures included TMS-evoked potentials (a measure of cortical excitability) at both the L-DLPFC (treatment site) and vertex Cz (control site), and sgACC source-localized activation.

**Results:** Active SNT progressively reduced cortical excitability specifically at the treatment site, with significant reduction observed by day 3 (N45 [early negative component at ∼45ms]: -27.9%, p<0.01; P200 [late positive component at ∼200ms]: -39.8%, p<0.001), while no changes occurred at the control site (Cz) in either group (all p>0.05). This spatial specificity was confirmed by direct site comparisons showing significant reductions only at the L-DLPFC target (t(22)=-3.8, p<0.001). Additionally, active SNT significantly reduced sgACC responses to TMS across sessions (F(13,222)=4.93, p<0.001), with effects persisting at 1-month follow-up. Critically, baseline sgACC activation predicted treatment response (r=-0.67, p=0.023), with higher baseline activation associated with greater clinical improvement.

**Conclusions:** SNT produces progressive, site-specific modulation of TMS-evoked potentials (cortical excitability) at the targeted L-DLPFC and selective reduction in sgACC activation, with neurophysiological changes mirroring clinical improvement. The spatial specificity of effects confirms target-specific neuroplasticity. Baseline sgACC activity and changes in cortical excitability early in treatment represent promising biomarkers for personalizing TMS treatment for depression.

## INTRODUCTION

Major depressive disorder (MDD) affects over 322 million people worldwide and represents the leading cause of disability globally (1, 2). Approximately 30% of MDD patients develop treatment-resistant depression (TRD), a condition where traditional antidepressant treatments like medication and therapy fail to produce meaningful clinical improvement (1). For patients with TRD, non-invasive brain stimulation techniques such as repetitive transcranial magnetic stimulation (rTMS) targeting the left dorsolateral prefrontal cortex (L-DLPFC) represent a promising alternative treatment approach (3).

Stanford Neuromodulation Therapy (SNT) is a high-dose, accelerated intermittent theta burst stimulation (iTBS) protocol that uses functional connectivity–guided targeting. In a randomized sham-controlled trial, SNT achieved a 62% mean reduction in depression severity after five days, with a large effect size (Cohen’s d = 1.71) over a much shorter timeframe than conventional rTMS protocols requiring several weeks of treatment (4). Despite these impressive clinical outcomes, the neurophysiological mechanisms underlying SNT’s therapeutic effects still require further investigation. Recent studies have begun to elucidate the neural correlates of SNT’s antidepressant effects using resting-state functional MRI (fMRI). Batail et al. (2023) reported modulation of functional connectivity involving the default mode network, amygdala, salience network, and striatum following SNT treatment (5). Similarly, Gajawelli et al. (2024) observed increased anti-correlation between the L-DLPFC and the default mode network following SNT, with changes sustained at one-month follow-up (6). However, the limited temporal resolution of fMRI constrains mechanistic insight into fast-acting neurophysiological changes induced by SNT.

Transcranial magnetic stimulation combined with electroencephalography (TMS-EEG) offers millisecond-level resolution of cortical excitability and connectivity (7–11). TMS-evoked potentials (TEPs), including the N45 (negative deflection at ∼45ms), N100 (negative deflection at ∼100ms), and P200 (positive deflection at ∼200ms) components, index excitation/inhibition balance and have shown abnormalities in MDD that normalize with treatment (12–14). Yet, no study has examined the progressive, day-by-day neurophysiological changes that occur during rapid, high-dose SNT treatment, leaving a critical gap in our understanding of how accelerated approaches achieve their rapid and robust clinical effects.

In this study, we used single-pulse TMS-EEG to track neurophysiological changes during the 5-day SNT protocol in patients with TRD, leveraging data from the original double-blind randomized trial. TMS-EEG was recorded at baseline, before and after each treatment day, and at one-month follow-up (n=24; 12 active, 12 sham). We assessed global TEP components (N45, N100, P200), Global Mean Field Amplitude, and source-localized sgACC responses to characterize cortical excitability changes both globally (via global TEP components and GMFA) and at specific brain regions (via source-localized sgACC responses). Based on evidence that iTBS induces long-term potentiation-like plasticity and modulates cortical excitability (15–17), and that high-dose protocols may produce cumulative plastic changes (18, 19), we hypothesized that SNT would induce progressive reductions in cortical reactivity and that these changes would correlate with clinical improvement and persist at post-treatment follow-up.

## METHODS

### Participants

Twenty-four participants (12 active, 12 sham) were included from a double-blind, randomized, sham-controlled trial of Stanford Neuromodulation Therapy (SNT) for treatment-resistant depression (TRD) (ClinicalTrials.gov NCT03068715) (4). All met Maudsley criteria for TRD and had moderate-to-severe depression, defined by the Montgomery-Åsberg Depression Rating Scale (MADRS) scores ≥20 (20). All were TMS-naïve and had failed at least one adequate antidepressant trial. Exclusion criteria included history of seizures, recent substance use disorder, psychosis, unstable medical illness, active suicidal intent, and contraindications to TMS or MRI. Written informed consent was obtained from all participants. The study was approved by the Stanford University Institutional Review Board. The study followed CONSORT guidelines for randomized controlled trials.

### Stanford Neuromodulation Therapy (SNT)

SNT consisted of 5 consecutive days of high-dose intermittent theta burst stimulation (iTBS), with 10 sessions per day spaced by 50-minute intervals. Stimulation targeted the left dorsolateral prefrontal cortex (L-DLPFC) region showing maximal anticorrelation with the subgenual anterior cingulate cortex (sgACC), identified via resting-state fMRI. Each iTBS session delivered 1800 pulses (triplets at 50 Hz repeated at 5 Hz over 2 seconds, 8-second inter-train interval), totaling 90,000 pulses. Stimulation was applied at 90% of resting motor threshold using a MagPro X100 stimulator (MagVenture) with a Cool-B70 coil. Resting motor threshold was determined using neuronavigated single-pulse TMS over the primary motor cortex with depth correction modeling to account for scalp-to-cortex distance (21). Sham stimulation followed the identical protocol but with the coil held at 90° to the scalp to mimic auditory and somatosensory input without inducing cortical activation. Both participants and raters were blinded to treatment conditions.

### TMS-EEG Data Acquisition and Processing

Single-pulse TMS-EEG recordings were acquired at baseline, before and after the 9th treatment session on each of the five treatment days (days 1-5), and at 1-month follow-up (Figure 1A). The 9th session was chosen to capture neurophysiological changes after substantial daily treatment exposure while accommodating EEG setup time constraints. Pulses were delivered to the individualized L-DLPFC target at 120% of resting motor threshold. EEG was recorded using a 64-channel BrainAmp DC system (Brain Products GmbH), digitized at 5000 Hz, with electrodes referenced to FCz and impedances kept below 5 kΩ. Each session included approximately 150 pulses with jittered inter-trial intervals of 4–6 seconds.

**Figure 1.**
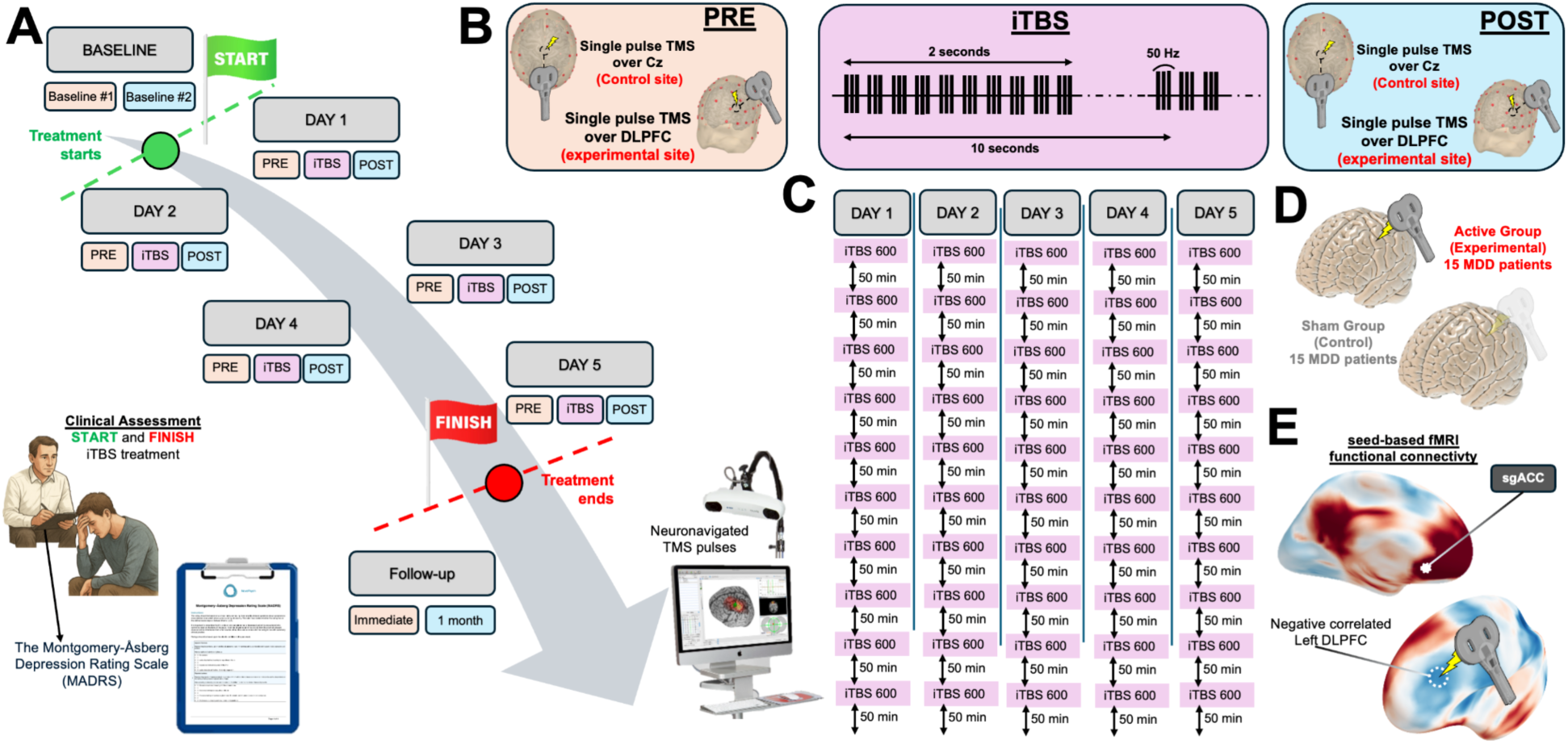
Stanford Neuromodulation Therapy (SNT) Protocol and Experimental Design. **A)** Study timeline illustrating the 5-day accelerated treatment protocol with baseline assessment, daily interventions, and follow-up evaluations. The treatment was Stanford Neuromodulation Therapy (SNT). Each treatment day includes pre-intermittent Theta Burst Stimulation (iTBS), and post-iTBS assessment. Pre-iTBS and post-iTBS consisted of single-pulse TMS targeting either the left DLPFC (treatment site) or Cz (control site) for neurophysiological measurements before and after each daily treatment session. Clinical assessments (bottom left) were performed before treatment (baseline), immediately post-treatment, and at 1-month follow-up using the Montgomery-Åsberg Depression Rating Scale (MADRS). **B)** Single-pulse TMS procedure targeting either the left DLPFC (treatment site) or the vertex of the scalp (Cz - control site) for comparison The single-pulse TMS were acquired every day both pre (orange) and post (light blue) the iTBS protocol (purple). The iTBS protocol consisted of 50 Hz triplet bursts delivered every 200ms (5 Hz) in 2-second trains followed by 8-second intervals over 10-second cycles. C) Schematic overview of the SNT protocol which consisted of 10 sessions per day of 600 pulses per session for a total of 6,000 pulses per day (ISI=intersession interval). **D)** Study groups comprising 12 patients with major depressive disorder (MDD) in the active treatment group and 12 patients in the sham control group. **E)** TMS targets were individualized using seed-based functional MRI connectivity analysis. The left DLPFC region showing the strongest anti-correlation with the subgenual anterior cingulate cortex (sgACC) was selected as the stimulation target.

Data preprocessing was performed using the AARATEP pipeline (v2.0), optimized for TMS-EEG artifact removal (22). Preprocessing included artifact interpolation, ICA-based rejection using ICLabel and TMS-specific criteria, and re-referencing to the average of all channels. Further technical details are provided in the Supplementary Methods.

### Analytical Approach

To ensure that consistent neurophysiological components were being measured across sessions, TEP morphological consistency was evaluated using cosine similarity (23) measures within and between participants across full TEP epochs (63 channels × 385 timepoints). Although it is difficult to evaluate TEP reliability during an active treatment protocol that is designed to induce neuroplastic changes, we assessed TEP morphological stability across days during SNT treatment using intraclass correlation coefficients (ICCs). This analysis evaluated whether the overall shape and topographic pattern of TEPs remained consistent across sessions, allowing us to distinguish systematic amplitude changes (reflecting treatment effects) from random variability or morphological drift. ICCs were computed from all measurement sessions, with values above 0.7 considered excellent. Full TEP morphological consistency metrics are available in the Supplementary Methods.

In order to examine the global cortical response to SNT, we evaluated global TMS-evoked potentials (TEPs). Global mean field amplitude (GMFA), calculated as the spatial standard deviation of voltage across channels, was analyzed across three canonical post-stimulation windows: 20–80 ms (termed the N45), 80– 150 ms (termed the N100), and 150–250 ms (termed the P200), defined a priori (24–26). The area under the curve (AUC) for each GMFA window was computed globally across all 64 channels and normalized to the pre-treatment baseline session (Session AUC / Pre-treatment Baseline AUC × 100). Linear mixed-effects models (LME) were used to examine group × time effects across 14 sessions to account for missing data, with Bonferroni correction for multiple comparisons (α = 0.0036).

Early local responses can be assessed with early local TEPs (EL-TEPs), which have been shown to be reliable and spatially specific markers of local cortical excitability (10, 11, 27). For early local TEP analysis, we sought to determine whether treatment effects were spatially restricted to the stimulated site or represent global network reverberation responses. Electrodes closest to the stimulation sites were identified based on Euclidean distance calculations. For both the left DLPFC (treatment site) and the vertex Cz (control site), the five nearest electrodes were selected based on Euclidean distance. Early local TEPs were computed as the spatial standard deviation of voltage across these electrode clusters within the [20-80 ms] time window for each stimulation site. Linear mixed-effects models (LME) were used to examine group × time effects across 14 sessions to account for missing data, with Bonferroni correction for multiple comparisons (α = 0.0036).

To examine topographic changes in TEPs amplitude patterns, we performed a two-stage permutation testing approach. First, within-subject permutation tests (1000 iterations) with cluster-based correction compared post-treatment versus baseline waveforms for each participant separately. Second, between-group permutation tests (1000 iterations) with cluster-based correction compared the magnitude of within-subject changes between active and sham groups across all channels and timepoints. This approach yielded topographic z-score maps identifying significant spatial-temporal clusters where active SNT produced greater changes than sham treatment.

To quantify how individual participants’ neurophysiological changes matched the group-level spatial patterns, we applied singular value decomposition (SVD) to extract dominant topographic components from each participant’s data (28). For each significant cluster identified in the spatiotemporal permutation analysis described above, we extracted 40-ms windows centered on peak latencies (∼49ms, ∼137ms, ∼242ms) from individual participant data. SVD was applied to these windows, and the first spatial component (left singular vector) was extracted as the dominant topographic pattern. We then computed cosine similarity between each participant’s dominant spatial pattern and the corresponding group-level difference map. Temporal strength was quantified using the AUC of the first temporal component (right singular vector).

Euclidean distance from baseline topographical patterns was calculated for each session to quantify the magnitude of neurophysiological change across treatment days. For early versus late adaptor classification, participants showing ≥25% reduction in N45 amplitude by day 3 were classified as early “adaptors”, while those reaching this threshold after day 3 were classified as late “adaptors”. Associations between adaptation timing and clinical response were examined using Pearson correlations.

For sgACC-targeted analysis, source-level activity was extracted from an ROI centered at MNI coordinates [4, 21, –8], derived from meta-analyses of depression-relevant circuitry (29, 30). Activation time courses were normalized to the pre-treatment baseline session and analyzed using linear mixed-effects models (group × time) to account for missing data, with Bonferroni-corrected post-hoc testing. Source-localized trials were excluded if the estimated current dipole strength fell below 10% of the maximum observed activation across all timepoints, indicating insufficient signal-to-noise ratio for reliable source estimation. Early sgACC changes were defined as the average change from pre-treatment baseline across days 1-3 for both active and sham SNT groups. Associations between early sgACC changes and clinical response (MADRS reduction at post-treatment and 1-month follow-up) were examined using Pearson correlations separately for active and sham groups.

Further technical and statistical details, including preprocessing, denoising thresholds, ROI definition, and SVD parameterization, are reported in the Supplementary Methods.

## RESULTS

### Spatio-temporal multi-session morphological consistency of TEPs

We first asked whether TMS-evoked potentials provide stable, individual-specific morphological signatures that could reliably track neuroplastic changes across the intensive 5-day SNT protocol. Visual inspection revealed that global TEPs showed consistent morphology across five consecutive treatment days, with stable topographies of canonical components (P15, N25, N45, N100, P200; Figure 2A). Quantitative cosine similarity analysis confirmed these observations across multiple dimensions. Within-subject cosine similarity maps demonstrated high consistency (yellow regions), while between-subject comparisons revealed lower similarity (blue regions), indicating unique individual neurophysiological signatures (Figure 2B). Session-by-session similarity analysis across all 14 sessions revealed excellent consistency (mean cosine similarity = 0.76 ± 0.05), particularly between adjacent days (Figure 2C). Critically, TEPs demonstrated strong individual specificity: within-subject similarity (0.733 ± 0.104) significantly exceeded between-subject similarity (0.403 ± 0.077; t(20) = 20.81, p < 0.001) (Figure 2D), confirming that each participant exhibits a unique, stable neurophysiological signature that is highly consistent within individuals but distinct between individuals. This individual specificity suggests that TEPs may potentially serve as reliable neurophysiological markers for tracking systematic changes across intensive treatment protocols (23, 31).

**Figure 2.**
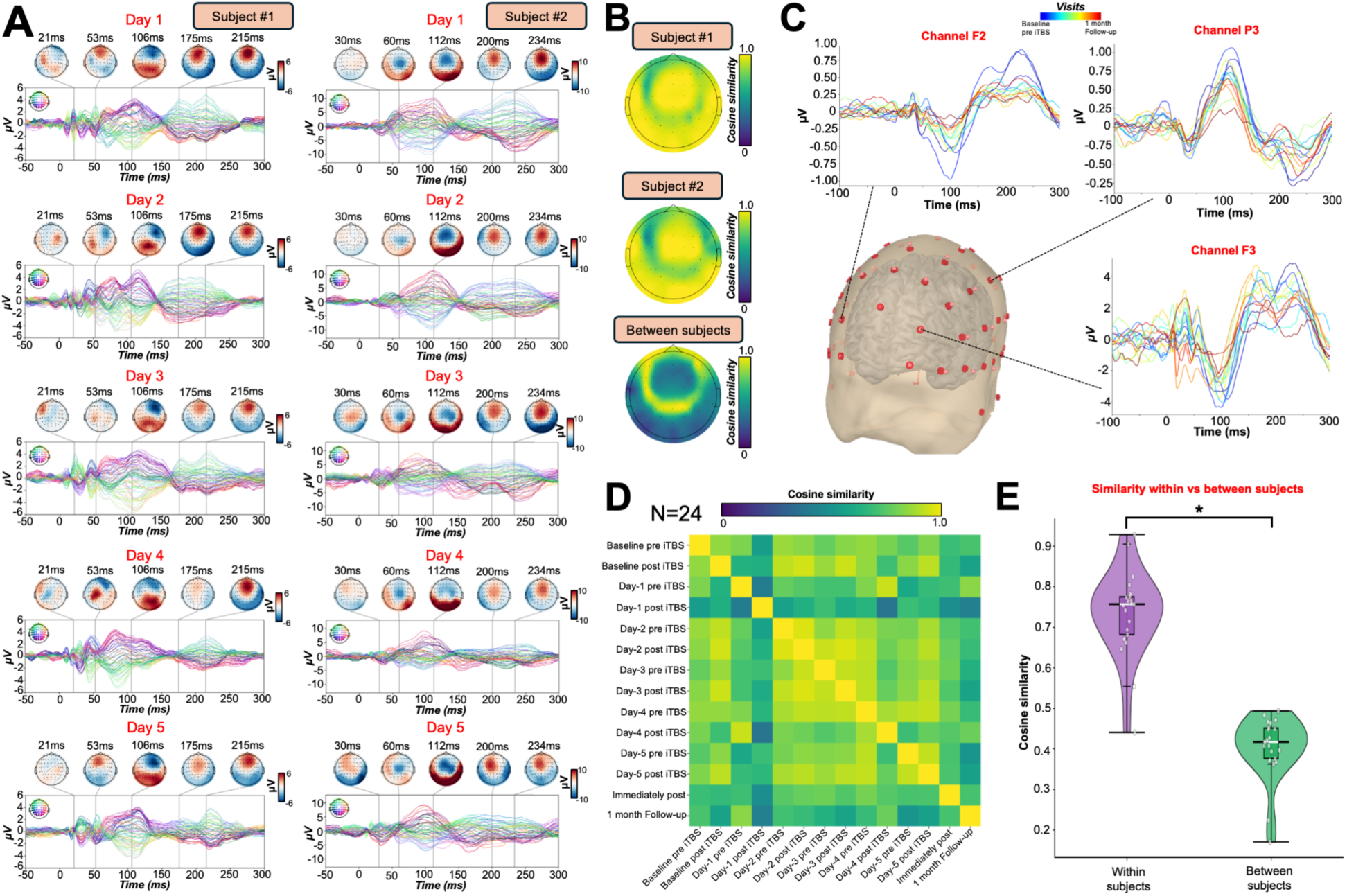
TEP morphological consistency across multiple visits. **A)** TEP topoplot for two representative participants shown across five consecutive pre-treatment days, with topographic maps of TEP components at specific latencies demonstrating consistent spatial distribution of activity patterns across treatment days. This shows that TEP topographies remain stable across multiple sessions, validating the use of TEPs as stable neurophysiological markers. **B)** Cosine similarity maps comparing TEP patterns within and between participants. Top panels show within-subject similarity for two representative participants across sessions (high similarity indicated by yellow regions), while the bottom panel shows between-subject similarity (lower similarity indicated by blue regions), demonstrating individual-specific neurophysiological signatures. **C)** Morphological consistency of TEP waveforms recorded from three channels: F3 (stimulation target site), P3 (same hemisphere, implicated in the same network), and F2 (contralateral hemisphere) across all sessions for one representative subject, color-coded by visit timing from baseline (blue) to 1-month follow-up (red). TEP morphology is preserved across all recording sites and sessions, confirming that treatment-related amplitude changes reflect genuine neuroplastic modifications. **D)** Session-by-session similarity matrix showing the cosine similarity between TEP patterns across all 14 sessions for the same representative subject. Higher values (yellow) indicate greater similarity between sessions. High cosine similarity values (mean = 0.76 ± 0.05) demonstrate excellent session-to-session consistency, particularly between adjacent days. **E)** Violin plot comparing within-subject similarity (left, mean = 0.733 ± 0.104) versus between-subject similarity (right, mean = 0.403 ± 0.077) across the entire sample (N=24), demonstrating that each participant exhibits a unique, stable neurophysiological signature that is highly consistent within individuals but distinct between individuals. This significant difference (t(20) = 20.81, p < 0.001) confirms that TEPs provide subject-specific measures of cortical excitability that are stable over time.

### Progressive Modulation of TMS-Evoked Neural Responses

We next asked whether SNT produces progressive changes in cortical excitability that distinguish active from sham treatment. We observed differential trajectories of cortical reactivity across treatment sessions in active versus sham groups at the treatment site (Figure 3A). LME models on normalized GMFA values for the N45 component (20–80 ms), with TIME (14 timepoints) as a within-subjects factor and GROUP (active vs. sham) as a between-subjects factor, revealed a significant GROUP × TIME interaction (F(13,286) = 4.23, p < 0.001, η² = 0.10), as well as main effects of GROUP (F(1,22) = 8.76, p = 0.007, η² = 0.19) and TIME (F(13,286) = 3.92, p < 0.001, η² = 0.09). In the active group, N45 GMFA decreased steadily from baseline (100%) to 72.1% ± 0.4% by day 3 post-iTBS and 60.2 ± 0.4% by day 5 post-iTBS, with effects persisting immediately post-treatment (66.4 ± 0.5%) and partially rebounding at 1-month follow-up (72.1 ± 0.4%) (Figure 3A). The sham group showed no systematic change (98.3–101.4%). Post-hoc tests (Bonferroni-corrected) revealed significant between-group differences starting from day 3 pre-iTBS (p < 0.01). For later components, LME revealed a non-significant GROUP × TIME interaction for N100 (F(13,286) = 1.76, p = 0.21, η² = 0.07) and a marginal effect for P200 (F(13,286) = 1.82, p = 0.042, η² = 0.08), but no significant post-hoc differences after correction (Figure 3A). In contrast, after applying single-pulse TMS at the control site (Cz), no significant changes were observed for any TEP component in either active or sham groups across treatment sessions (all p > 0.05) (Figure 3B). This spatial specificity was further confirmed by direct comparisons showing significant reductions at the treatment site for the N45 component (t(22) = -3.8, p < 0.001) at both immediate post-treatment and 1-month follow-up, while no changes were observed at the control site or for other TEP components (all p > 0.05) (Figure 3C). Early local TEP analysis corroborated these findings, demonstrating significant suppression of evoked responses measured specifically at the treatment site in the active group (F(13,156) = 3.2, p < 0.001), while local responses at the control site remained stable across both groups (F(13,156) = 0.8, p = 0.65) (Figure 3D).

**Figure 3.**
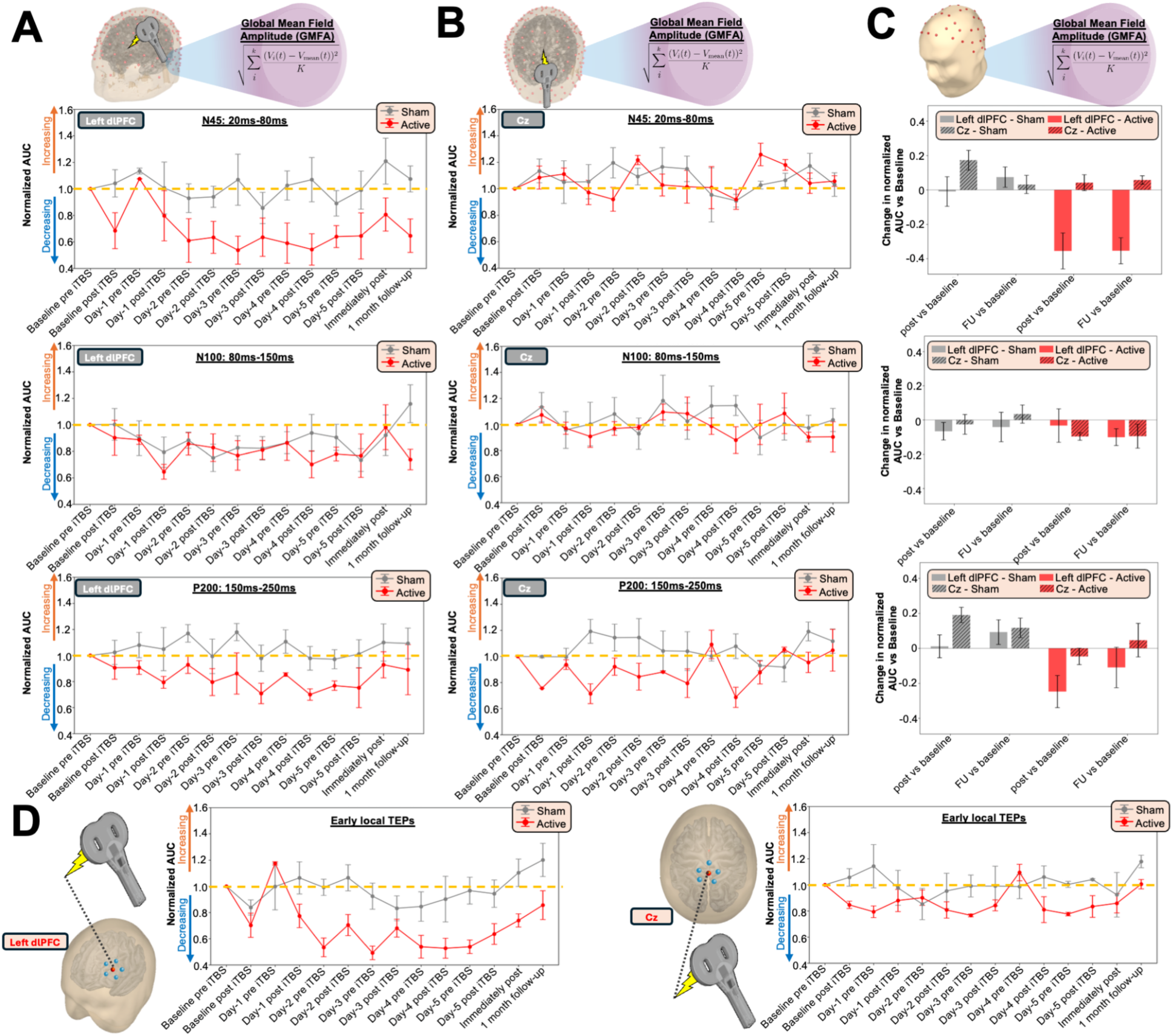
A) Progressive Modulation of TMS-Evoked Neural Responses Following Five Days of SNT Protocol. **A)** Normalized area under the curve (AUC) of the global mean field amplitude (GMFA) for three time windows corresponding to established TMS-evoked potential peaks (N45: 20ms-80ms; N100: 80ms-150ms; P200: 150ms-250ms) recorded from the left DLPFC (treatment site). Data are normalized to individual baseline values. The active SNT group (red) shows a progressive decrease in neural response amplitude for the early (N45: 20-80ms) and late (P200: 150-250ms) components across treatment days, while showing no significant changes in the intermediate component (N100: 80-150ms). The sham group (grey) maintains relatively stable responses across all three TEP components. **B)** Normalized AUC of GMFA for the same three TEP components recorded from the vertex Cz (control site). Both active (red) and sham (grey) groups show no changes across treatment sessions, demonstrating the site-specificity of the neurophysiological effects observed at the treatment site. **C)** Bar plots comparing changes in normalized AUC between treatment (left DLPFC) and control (Cz) sites for both active and sham groups at immediate post-treatment and 1-month follow-up timepoints. The active group shows significant reductions specifically at the treatment site (red bars) for the early (N45: 20-80ms) and late (P200: 150-250ms) components, with no changes for the intermediate component (N100: 80-150ms), while showing no significant changes at the control site. The sham group shows no significant changes at either site (grey bars), confirming the treatment-specific and site-specific nature of the neurophysiological modulation. **D)** Early local TEPs showing detailed waveforms and time courses for the treatment site (left DLPFC, left panel) and control site (Cz, right panel). The treatment site demonstrates clear suppression of evoked responses in the active group (red) compared to sham (grey), while the control site shows stable responses across both groups, further confirming the spatial selectivity of SNT-induced neuroplasticity.

### Spatiotemporal Dynamics of Neurophysiological Change and Clinical Relevance

We characterized the evolution of cortical plasticity during SNT by combining spatiotemporal cluster permutation testing, singular value decomposition (SVD), and trajectory tracking. First, channel-wise cluster permutation tests compared baseline and post-treatment TEPs across all electrodes and timepoints, revealing three significant windows of treatment-related change in the active group (Figure 4A). The earliest cluster, peaking at ∼49 ms (N45), was centered over frontocentral electrodes (peak z = –3.42, *p* < 0.01, cluster-corrected). A second cluster at ∼137 ms (N100) emerged over central-parietal regions (z = – 3.86, *p* < 0.005), and a third at ∼242 ms (P200) was widely distributed over frontoparietal cortex (z = –4.15, *p* < 0.001). No significant changes were observed in the sham group. This spatial progression—from localized early effects near the stimulation site to later widespread changes, suggests that SNT may induce plasticity that propagates through connected cortical networks.

**Figure 4.**
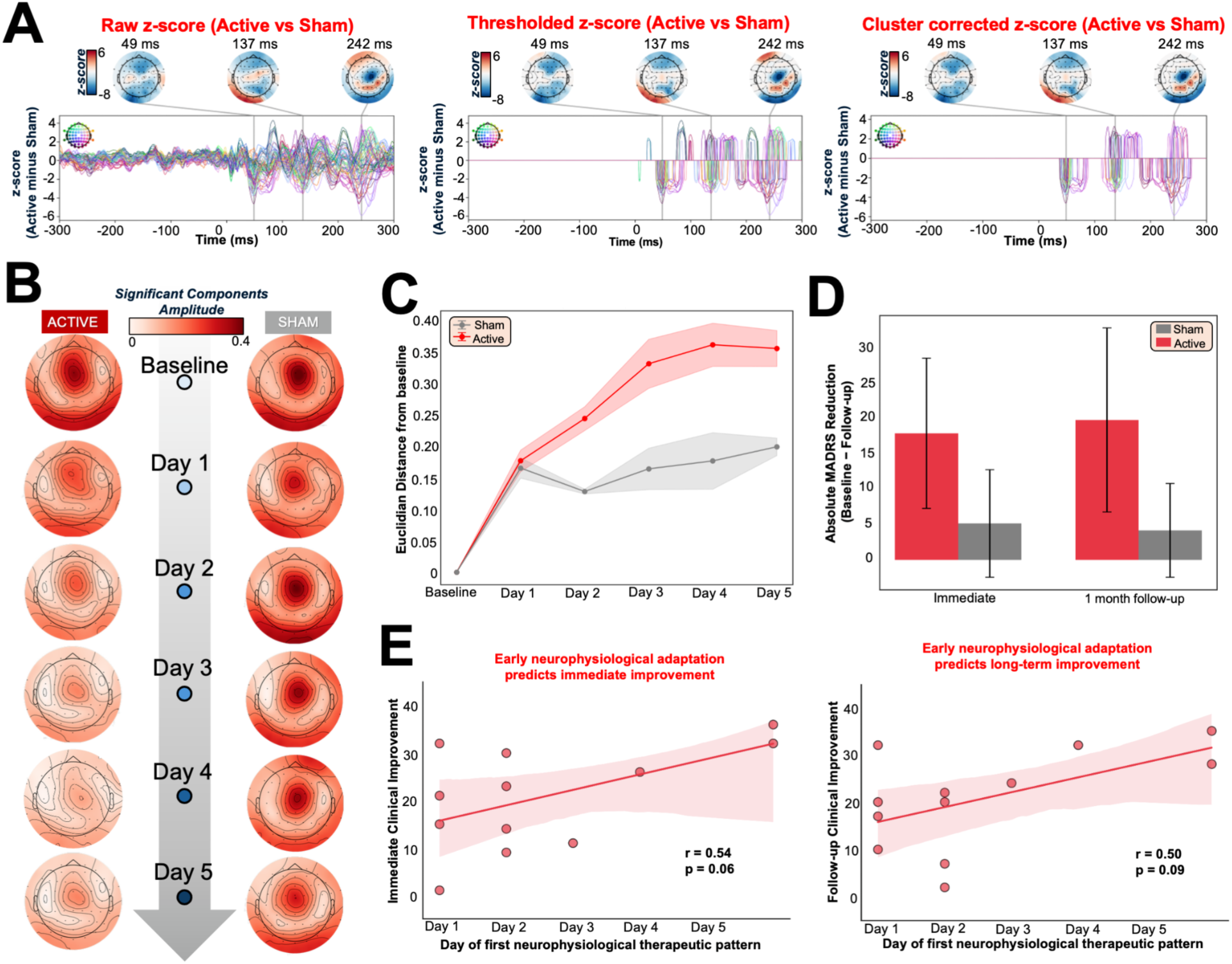
Topographical Neurophysiological Changes and Early Adaptation in SNT Treatment for Major Depression. **A)** Spatiotemporal cluster-based permutation analysis comparing TMS-evoked responses between first and last treatment visits. Three significant time windows were identified at approximately 49ms, 137ms, and 242ms post-TMS pulse. The analysis shows topographic z-score maps representing statistical differences in neural responses, with negative values (blue) indicating reduced amplitude in the active group compared to sham. The progression from focal frontal effects (49ms) to more distributed frontoparietal changes (137ms, 242ms) suggests intervention-induced plasticity spreading across interconnected networks. **B)** Evolution of topographical patterns identified through singular value decomposition (SVD) analysis of the three significant time windows identified in panel A. Topoplots illustrate the progressive decrease in neural response amplitude (represented by color intensity) across treatment days (baseline through day 5) in the active group (left column), while the sham group (right column) maintains consistent response patterns. Blue circles indicate the progression of treatment days. This analysis demonstrates the cumulative effect of repeated SNT protocol sessions on cortical excitability specifically in the active treatment group, with effects persisting through the 1-month follow-up period. **C)** Euclidean distance from baseline topographical patterns, quantifying the magnitude of neurophysiological change across treatment days corresponding to the SVD-derived patterns shown in Panel B. For each day, the topographical patterns were averaged across pre- and post-iTBS sessions (baseline through day 5), and Euclidean distance was calculated between each day’s pattern and the baseline topography. The active group (red) shows progressive increases in distance from baseline, indicating cumulative topographical reorganization, while the sham group (grey) maintains minimal deviation from baseline patterns throughout treatment. **D)** Clinical improvements (absolute changes) on the MADRS depression scale were more pronounced in the active group both immediately after treatment and at 1-month follow-up. **E)** Relationship between the day of first significant neurophysiological change and clinical improvement measured by MADRS reduction at immediate post-treatment (left) and 1-month follow-up (right). For each patient, we identified the earliest day showing meaningful neurophysiological change from baseline in the therapeutic patterns identified in panels A and B, using optimized thresholds and multiple detection criteria to capture individual adaptation patterns. The scatter plots show the relationship between this timing measure and clinical outcomes, with earlier neurophysiological changes potentially associated with better treatment response. While trends are visible in both timepoints (immediate: r = 0.54, p = 0.06; 1-month: r = 0.50, p = 0.09), these relationships do not reach statistical significance. The analysis represents an exploratory approach to identify potential biomarkers of treatment response timing.

To probe individual-level expression of these group-level effects, we applied SVD analysis to assess how closely each subject’s neurophysiological changes aligned with the group-defined spatiotemporal signatures of SNT-induced plasticity (see Methods for details).

The temporal amplitude of these subject-specific components was then tracked across the 14-session protocol. In the active group, SVD-derived amplitudes declined to ∼60% of baseline by day 5, with partial recovery at 1-month follow-up. The sham group showed no systematic change (Figure 4B). To quantify treatment-related reorganization, we calculated the Euclidean distance between each day’s topography and the baseline pattern. In the active group, this distance increased progressively over time—peaking at days 3–5—while remaining stable in sham participants (GROUP × TIME interaction: F(13,286) = 3.21, p < 0.001; Figure 4C). These metrics confirmed the cumulative and treatment-specific nature of the observed neurophysiological plasticity.

We next asked how individual neurophysiological trajectories related to group-level patterns and clinical response. Clinical improvements were significantly greater in the active group compared to sham, both at post-treatment (Δ = –7.5, t(44) = –4.02, p = 0.0002) and at 1-month follow-up (Δ = –6.0, t(44) = –3.03, p = 0.004), confirming the sustained antidepressant effect of SNT (Figure 4D).

Based on these TEPs trajectories, we identified early adaptors as participants who reached a ≥25% reduction in SVD amplitude by day 3 (n = 10), and late adaptors as those crossing this threshold only afterward (n = 2). Earlier adaptation was associated with greater clinical improvement, with moderate correlations between day-of-adaptation and MADRS reduction both post-treatment (r = 0.54, p = 0.07) and at 1-month follow-up (r = 0.50, p = 0.10; Figure 4E). Although these trends did not reach significance, they suggest that the timing of neurophysiological engagement may serve as a predictor of clinical response.

Together, these results support a model in which SNT induces progressive and spatially expanding cortical plasticity. Early alignment with treatment-induced topographies may reflect a subject’s capacity to engage key neural circuits, offering a potential avenue for biomarker-driven response monitoring and individualized protocol refinement.

### sgACC Suppression and Its Relationship to Treatment Response

Next, we investigated whether SNT modulates sgACC activity, given its central role in depression and established functional connectivity with the L-DLPFC (32–35). Source-localized analysis of sgACC activity (MNI coordinates [4, 21, –8]) revealed significant treatment-specific modulation over the SNT protocol. LME modeling demonstrated a significant GROUP × TIME interaction (F(13,222) = 4.93, p < 0.001), indicating divergent neurophysiological trajectories between active and sham conditions (Figure 5A). The active group exhibited immediate and sustained sgACC suppression beginning from the first post-iTBS session, with activity consistently below normalized baseline levels, while the sham group maintained stable activation throughout treatment.

**Figure 5.**
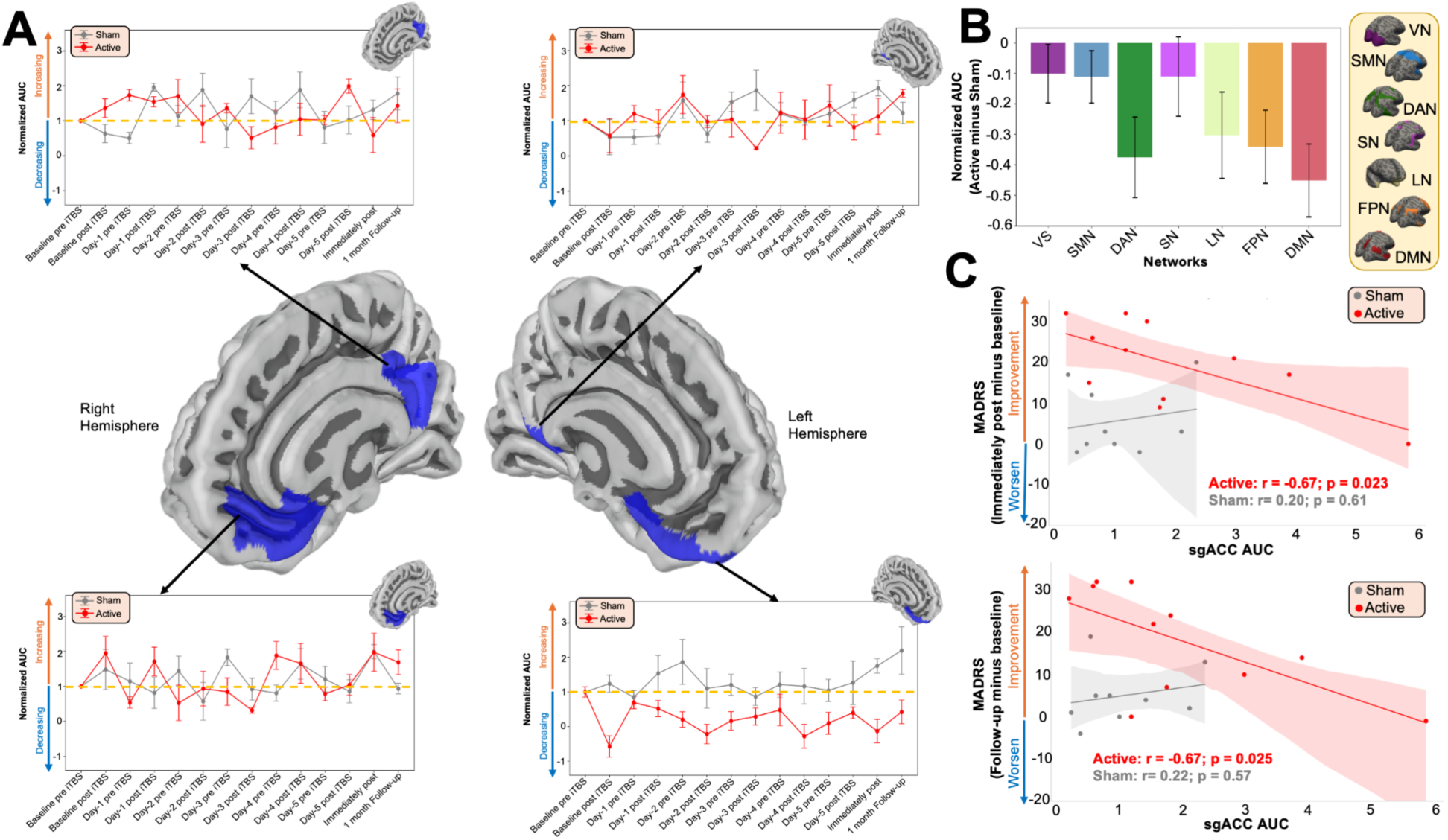
Regional and Network-Specific Neural Response to SNT Treatment and Predictive Biomarkers. **A)** Changes in normalized source-localized area under the curve (AUC) across treatment sessions for the sgACC (target region of SNT treatment) and control regions matched for depth from scalp and distance from the therapeutic stimulation site (left DLPFC). Four panels show normalized AUC for these brain areas, with the active SNT group (red) and sham group (grey) displaying distinct trajectories. Significant reduction was observed only in the sgACC following SNT treatment, while control regions show no significant changes, demonstrating the specificity of therapeutic modulation to the intended target circuit. **B)** Network-level analysis showing normalized AUC changes for major brain networks including Visual Network (VN), Salience/Somatomotor Network (SMN), Dorsal Attention Network (DAN), Sensorimotor Network (SN), Limbic Network (LN), Frontoparietal Network (FPN), and Default Mode Network (DMN). Bar plots demonstrate that the active group shows significant reductions in most networks (particularly DAN, LN, FPN, and DMN), while the sham group shows no significant changes across all networks. The widespread network modulation suggests that SNT produces broad-scale neural reorganization beyond the primary sgACC target. **C)** Predictive value of baseline sgACC activity for clinical outcomes. Scatter plots show the relationship between baseline sgACC AUC and MADRS improvement at immediate post-treatment (top) and 1-month follow-up (bottom). In the active group (red), higher baseline sgACC activity significantly predicts greater clinical improvement at both timepoints (immediate: r = -0.67, p = 0.023; follow-up: r = -0.67, p = 0.025), while the sham group (grey) shows no significant relationship. This suggests that baseline sgACC activity serves as a specific biomarker for SNT responsiveness, with patients showing higher baseline sgACC activation having greater capacity for therapeutic modulation.

Bonferroni-corrected post-hoc comparisons confirmed significantly reduced sgACC activation in the active group at multiple critical timepoints: day 1 post-iTBS (Δ = -1.02, p < 0.01), day 3 post-iTBS (Δ = -1.65, p < 0.001), immediate post-treatment (Δ = -1.88, p < 0.001), and crucially, at 1-month follow-up (Δ = -1.77, p < 0.001), demonstrating persistent modulation of this depression-relevant circuit. To control for potential depth-related artifacts inherent to EEG source reconstruction, we analyzed matched control regions at equivalent cortical depths and distance both ipsilaterally and contralaterally to the sgACC. These control regions showed no significant GROUP × TIME interactions (all F < 1.2, all p > 0.25) in either treatment condition, confirming that the observed sgACC modulation might reflect genuine circuit-specific neuroplasticity.

Network-level analysis revealed widespread but selective modulation across major brain systems (Figure 5B). The active group demonstrated significant reductions in the Default Mode Network (Δ = -0.43, p < 0.001), Frontoparietal Network (Δ = -0.38, p < 0.01), Dorsal Attention Network (Δ = -0.35, p < 0.01), and Limbic Network (Δ = -0.32, p < 0.05), while Visual, Sensorimotor, and Salience networks remained unchanged. The sham group exhibited no significant network-level changes (all p > 0.05), indicating that SNT produces coordinated reorganization across depression-relevant circuits beyond the primary sgACC target.

Baseline sgACC activation showed a potential predictive relationship with treatment response in the active group (Figure 5C). Higher baseline sgACC activity was associated with greater MADRS improvement at immediate post-treatment (r = -0.67, p = 0.023) and at 1-month follow-up (r = -0.67, p = 0.025). This relationship was not observed in the sham group (immediate: r = 0.20, p = 0.613; follow-up: r = 0.18, p = 0.642), suggesting that baseline sgACC activity may index capacity for therapeutic modulation via the L-DLPFC–sgACC circuit, though replication in larger samples is needed to confirm this specificity.

These convergent findings demonstrate that SNT induces selective and persistent suppression of sgACC activation while simultaneously modulating functionally connected networks. The strong predictive value of baseline sgACC activation (r = -0.67) suggests this measure could serve as a precision medicine biomarker, enabling identification of patients most likely to benefit from SNT’s intensive protocol and potentially informing treatment allocation decisions in clinical practice.

## DISCUSSION

Using comprehensive daily TMS-EEG measurements during SNT in treatment-resistant depression, we identified several key neurophysiological signatures of therapeutic response. First, TMS-evoked potentials demonstrated excellent morphological consistency and individual specificity across the intensive 5-day protocol, validating their use as stable neurophysiological markers. Second, SNT induced progressive reductions in cortical excitability, with EL-TEPs amplitude decreasing by 27.9% by day 3 and reaching ∼40% suppression by treatment end. Third, spatiotemporal cluster analysis revealed three distinct windows of treatment-related change (49ms, 137ms, 242ms), with effects progressing from localized frontal regions to distributed frontoparietal networks. Fourth, early neurophysiological adaptation (≥25% reduction by day 3) was associated with superior clinical outcomes compared to late adaptation. Finally, source-localized analysis demonstrated selective and sustained suppression of sgACC activation throughout active treatment, with baseline sgACC activity strongly predicting clinical response (r=-0.67). Together, this work shows that SNT’s rapid antidepressant effects are mediated by cumulative neuroplastic changes in depression-relevant circuits serving as promising biomarkers for personalizing TMS treatment.

Our main finding reveals that progressive reduction in cortical excitability represents a key mechanism underlying SNT’s therapeutic effects. The active group showed a 27.9% reduction in EL-TEPs amplitude by day 3 and ∼40% by treatment end, while sham treatment produced no systematic changes (<5%). This temporal evolution aligns with SNT’s rapid antidepressant effects (4), indicating cumulative neuroplastic changes that build across treatment sessions rather than immediate effects. The progressive cortical suppression mirrors the timeline of clinical improvement reported in the original trial, where significant MADRS reductions emerged by day 3-4. This temporal concordance raises the possibility that neurophysiological changes may relate to the mechanisms underlying symptom improvement, though causality cannot be established. The magnitude of group-level EL-TEPs suppression (40% by day 5) showed a similar scale to the group-level mean reduction in depression severity (62%), though individual-level correlations between neurophysiological and clinical changes require further investigation. While the EL-TEP N45 component has been linked to inhibitory processes in prior TMS-EEG studies (12), the specific mechanisms underlying these amplitude reductions in our GMFA analysis require further investigation. The concurrent modulation of the P200 component, reflecting network-level propagation (36–38), suggests SNT affects both local cortical excitability and broader information processing networks. Neurophysiological changes persisted at 1-month follow-up with partial recovery (from 60% to 72% of baseline), indicating long-term depression (LTD)-like plasticity mechanisms. This partial recovery corresponds with the maintenance of clinical gains reported in the original trial, supporting a biological basis for SNT’s sustained efficacy. The persistence of these neuroplastic changes distinguishes SNT from standard rTMS protocols, which typically exhibit more transient effects (39), potentially explaining SNT’s superior and more durable therapeutic outcomes.

Our spatiotemporal analysis reveals that SNT induces selective circuit-level modifications rather than global cortical suppression. Cluster permutation testing identified three distinct windows of treatment-related change at 49ms, 137ms, and 242ms post-stimulation, each with unique topographical distributions. The earliest effects (49ms) were localized over frontocentral regions near the stimulation site, while later components showed progressively more distributed patterns extending to central-parietal (137ms) and frontoparietal networks (242ms). This temporal-spatial progression supports transsynaptic propagation through interconnected cortical networks (36, 40, 41), consistent with contemporary views of TMS as a network-level intervention that selectively engages circuits based on stimulation parameters (42). SVD analysis further demonstrated that individual participants’ neurophysiological changes aligned with these group-level patterns, with spatial components showing monotonic amplitude reduction to ∼60% of baseline by day 5. The progressive nature of these changes, quantified through Euclidean distance metrics, revealed cumulative cortical reorganization that was entirely absent in sham participants, confirming the treatment-specific nature of the observed plasticity. The timing of neurophysiological engagement emerged as a critical determinant of therapeutic success, with most active participants (10/12) demonstrating early adaptation within the first three treatment days, characterized by ≥25% reduction in cortical excitability markers. These early adaptors exhibited significantly stronger clinical improvements compared to the minority showing late adaptation, with moderate correlations between adaptation timing and MADRS reduction both post-treatment (r = 0.54) and at follow-up (r = 0.50). This pattern aligns with established principles linking early physiological changes to treatment response across psychiatric interventions (43, 44) and suggests the existence of critical periods of heightened neuroplasticity (45). SNT’s unique dosing strategy, featuring increased pulse density and optimized 50-minute inter-session intervals, likely capitalizes on spaced learning principles that enhance synaptic consolidation and plasticity (46, 47), differing fundamentally from conventional rTMS protocols using 24-hour spacing and potentially explaining both the rapid onset and enhanced durability of SNT-induced neuroplastic changes.

Finally, our EEG source-reconstruction analysis showed how sgACC might play a key mechanistic role in SNT’s therapeutic effects. Notably, the L-DLPFC stimulation site was specifically selected based on its maximal anti-correlation with the sgACC in each individual participant, making sgACC responses a particularly relevant measure of circuit-specific treatment effects. The active group demonstrated progressive sgACC suppression beginning from the first treatment session and persisting through 1-month follow-up, while sham participants maintained stable activity throughout. Given the well-established role of sgACC hyperactivity in depression pathophysiology (30, 33–35, 48), this selective normalization reflects direct therapeutic action on depression-relevant circuitry. Critically, baseline sgACC activity emerged as a strong predictor of clinical response (r = -0.67), independent of baseline symptom severity, suggesting that sgACC hyperactivity indexes neural circuit capacity for therapeutic modulation rather than simply reflecting current depression severity. This finding supports contemporary circuit-based models of depression, where functional connectivity to sgACC predicts TMS efficacy across different stimulation targets (33), and sgACC-guided targeting strategies improve clinical outcomes (34). Our results align with this framework, indicating that elevated baseline sgACC activity confers greater potential for therapeutic modulation via the L-DLPFC–sgACC circuit.

These neurophysiological signatures offer immediate pathways for transforming TMS practice from a ’trial-and-error’ to a ’precision medicine’ approach. For immediate clinical application, baseline sgACC activity assessment via task-free fMRI before SNT initiation could prioritize patients with higher activation levels most likely to respond. Day 3 neurophysiological monitoring could serve as an early checkpoint—patients without ≥25% N45 reduction may benefit from parameter adjustments in intensity, targeting, or session frequency before completing the full protocol. The 50-minute inter-session interval used in SNT may be important for enabling cumulative plasticity effects, consistent with evidence that spaced learning paradigms enhance synaptic consolidation (46, 47), though direct comparisons with alternative intervals are needed to confirm this hypothesis. Longer-term integration of portable EEG systems could enable routine biomarker-guided TMS monitoring across diverse clinical settings, maximizing therapeutic efficacy while minimizing treatment burden for predicted non-responders. While limitations include our modest sample size, particularly among late adaptors, and the lack of direct L-DLPFC–sgACC connectivity measurements, these findings provide a foundation for biomarker-driven TMS protocols that could significantly improve treatment outcomes for patients with treatment-resistant depression.

## CONCLUSION

NT induces progressive reduction of both global and local cortical excitability and sgACC activation, key neurophysiological changes underlying its therapeutic effects in treatment-resistant depression. The timing of neurophysiological adaptation and baseline sgACC activity emerge as potential predictive biomarkers of treatment response, with early neural adaptation and higher baseline sgACC activity associated with superior clinical outcomes. These neurophysiological signatures advance our understanding of accelerated TMS mechanisms and may inform future personalized approaches for treatment-resistant depression, potentially enabling the identification of optimal candidates and treatment parameters to maximize therapeutic efficacy.

## Data Availability

The datasets generated and analyzed during the current study are not publicly available due to patient privacy protections but are available from the corresponding author upon reasonable request and with appropriate ethical approval.

